# Higher-Order Dynamics Beyond Repolarization Alternans in Ex-Vivo Human Ventricles are Independent of the Restitution Properties

**DOI:** 10.1101/2023.08.16.23293853

**Authors:** Shahriar Iravanian, Ilija Uzelac, Anand D Shah, Mikael J Toye, Michael S. Lloyd, Michael A. Burke, Mani A Daneshmand, Tamer S Attia, J David Vega, Michael El-Chami, Faisal M. Merchant, Elizabeth M Cherry, Neal K. Bhatia, Flavio H. Fenton

## Abstract

**Background:** Repolarization alternans, defined as period-2 oscillation in the repolarization phase of the action potentials, provides a mechanistic link between cellular dynamics and ventricular fibrillation (VF). Theoretically, higher-order periodicities (e.g., periods 4, 6, 8,…) are expected but have minimal experimental evidence.

**Methods:** We studied explanted human hearts obtained from recipients of heart transplantation at the time of surgery. Optical mapping of the transmembrane potential was performed after staining the hearts with voltage-sensitive fluorescent dyes. Hearts were stimulated at an increasing rate until VF was induced. Signals recorded from the right ventricle endocardial surface prior to induction of VF and in the presence of 1:1 conduction were processed using the Principal Component Analysis and a combinatorial algorithm to detect and quantify higher-order dynamics. Results were correlated to the underlying electrophysiological characteristics as quantified by restitution curves and conduction velocity.

**Results:** A prominent and statistically significant global 1:4 peak (corresponding to period-4 dynamics) was seen in three of the six studied hearts. Local (pixel-wise) analysis revealed the spatially heterogeneous distribution of periods 4, 6, and 8, with the regional presence of periods greater than two in all the hearts. There was no significant correlation between the underlying restitution properties and the period of each pixel.

**Discussion:** We present evidence of higher-order periodicities and the co-existence of such regions with stable non-chaotic areas in ex-vivo human hearts. We infer from the independence of the period to the underlying restitution properties that the oscillation of the excitation-contraction coupling and calcium cycling mechanisms is the primary mechanism of higher-order dynamics. These higher-order regions may act as niduses of instability that can degenerate into chaotic fibrillation and may provide targets for substrate-based ablation of VF.

## Introduction

Malignant ventricular arrhythmias, including polymorphic ventricular tachycardia (VT) and ventricular fibrillation (VF), are the primary causes of sudden cardiac death in many patients. Therefore, understanding the dynamics of VF and its initiation is of utmost theoretical and practical importance.^1–3^

Current management of VF comprises membrane-active antiarrhythmic medications and implantable defibrillators.^4^ Ablation of monomorphic ventricular tachycardia is the cornerstone therapy for drug-refractory VT or VT storm. Ablation strategies include activation mapping during VT or substrate ablation, which focuses on locating the critical reentry circuit using the local electrophysiological properties, such as the peak-to-peak amplitude and the conduction velocity (e.g., in the form of isochrone crowding).^5–7^

Conversely, VF ablation is in its infancy. It is performed rarely and only in specific circumstances targeting triggers or fixed anatomical structures, such as the right ventricular outflow tract ablation in Brugada syndrome.^8–10^ However, due to the lack of detailed mechanistic insight into the initiation of VF, functional substrate ablation of VF is still not possible. For example, the concordant to discordant pathway to VF initiation is well-known and extensively studied but cannot guide ablation because of its spatially dynamic nature.^11–13^

The hope is that by moving beyond alternans to more complex dynamics, we may gain useful localizing information that can be used to find unstable substrates suitable for ablation. Therefore, one of the motivations of the current study is the hypothesis that *the electrical instability starts focally before expanding to the entire ventricles, and ablation of these areas has a stabilizing effect and may reduce the risk of VF*.

Before directly testing the feasibility of VF ablation, we need to verify the premise of the hypothesis, namely the existence and detectability of electrically unstable substrates before VF induction. We use complex repolarization dynamics as a surrogate for electrical instability. Action Potential Duration (APD) alternans is the beat-to-beat (period-2) oscillation in the APD, is the simplest quantifier of repolarization dynamics, and is the one used to describe discordant alternans.^14^ Most cardiac tissues exhibit APD alternans when stimulated at a sufficiently fast rate. APD alternans can even be detected in clinical settings as the microvolt T-wave alternans (TWA).^11^ However, classic alternans with period-2 results from the first bifurcation in a cascade of bifurcations with periods of 2, 4, 8… We anticipate that as the cycle length becomes shorter, higher-order periods (especially periods of powers-of-two) appear until the system transitions to chaos and hence VF. The intermediate stages in the period-doubling cascade in cardiac tissue have been experimentally elusive and are reported in only a few animal models,^15–17^ but not in human hearts.

In this paper, we are particularly interested in these higher-order periodicities. Specifically, using higher-order repolarization dynamics to measure electrical instability, we try to answer the following questions:

1. Do higher-order periods (> period-2) occur in (diseased) human hearts?
2. What is the spatial distribution of the higher-order areas (are they focal)?
3. Are the baseline electrophysiological characteristics predictive of the local dynamics?

## Methods

### Heart Harvesting

The study protocol was approved by the Emory University and Georgia Institute of Technology Institutional Review Boards (IRB). Patients consented to the research protocol before surgery. We obtained hearts from the recipients of orthotopic heart transplantation at Emory University Hospital. At the time of surgery, each patient was fully heparinized and placed on a cardiopulmonary bypass machine after circulatory arrest was induced with the infusion of cold cardioplegia solution. Then, the pericardium was opened, and the recipient’s heart was removed using the bicaval technique.

Within 5 minutes of heart harvesting, the explanted heart was perfused from the arteries with cold cardioplegia solution for 5 minutes and transported to the optical mapping lab. Once in the lab, the left main and right coronary arteries were cannulated, and the heart was perfused with warm (37C) oxygenated Tyrode’s solution until the return of the spontaneous contractions. The heart was placed in an imaging chamber and perfused for at least half an hour to recover from the cardioplegia. Before optical mapping, the cardiac motion was suppressed with the help of the myosin ATPase inhibitor (-)-Blebbistatin, at a concentration of 1.8 uM.^18^

### Optical Mapping

Optical mapping is one of the main tools to study complex arrhythmias.^19^ Staining arterially-perfused explanted whole heart or a segment with voltage-sensitive fluorescent dyes allows for non-contact mapping of the transmembrane potential with high spatial (sub-millimeter) and temporal (milliseconds) resolutions.

In this study, after the heart was prepared as above (arterially cannulated, perfused, and immobilized), it was stained with 1 mg of near-infrared voltage-sensitive dye JPW-6003 (also known as di-4-ANBDQPQ) dissolved in ethanol.^20,21^ The first voltage measurements were obtained from the epicardial surface as long as there was an acceptable imaging window (depending on the amount of fat across the epicardial surface). Afterward, the right ventricle free wall was removed, and its endocardial surface was imaged. We report on the results of transmembrane potential mapping from the endocardial surface of the right ventricles.

The tissue was excited using a deep red LED coupled with a 660/20 nm bandpass filter, and the emitted fluorescence was directed through a long pass 700 nm filter into an Electron-Multiplying-Charged-Coupled-Camera (EMCCD) camera. Image acquisition was performed at 500 Hz and with a resolution of 128 x 128 pixels.

### Study Protocol

The study protocol was organized as multiple restitution runs. In each run, the right ventricles were paced at progressively faster rates, starting at a pacing cycle length of ∼2000 ms and down to either the emergence of 2:1 block or induction of a reentrant arrhythmia (VT or VF). Optical signals were recorded from the endocardial surface in 20-40 seconds segments. We compared the signal recorded just before VT/VF induction (while still global 1:1 capture and conduction were present) to a control recording at 500 ms to detect higher-order periods.

### Signal Processing

The transmembrane potential data for each pixel is low-passed filtered by a cutoff of 50 Hz, and the output is normalized in the 0 to 1 range. Spatial smoothing is performed by applying a variational method (Supplement A).

Period-4 and higher-order dynamics are not distributed uniformly over the recording area.^15^ Instead, they are localized to a few regions. In response, we have developed two complementary processing pathways, one *global* to detect low-amplitude higher-order periodic signals from the whole imaging area and the other *local* to localize period-4 and higher at a pixel level.

Figure 1 explains the global analysis methodology (details in Supplement B). The output of the global analysis is a spectrogram, where the frequency is normalized to the pacing frequency, such that the 1:1 peak corresponds to the principal action potential propagation. We are mainly interested in the sub-harmonics of the 1:1 peak. The 1:2 peak (located at exactly half the driving frequency) is a sign of period-2 alternans. Similarly, the 1:4 peak is a marker of the period-4 oscillation in the repolarization phase.

**Figure 1.**
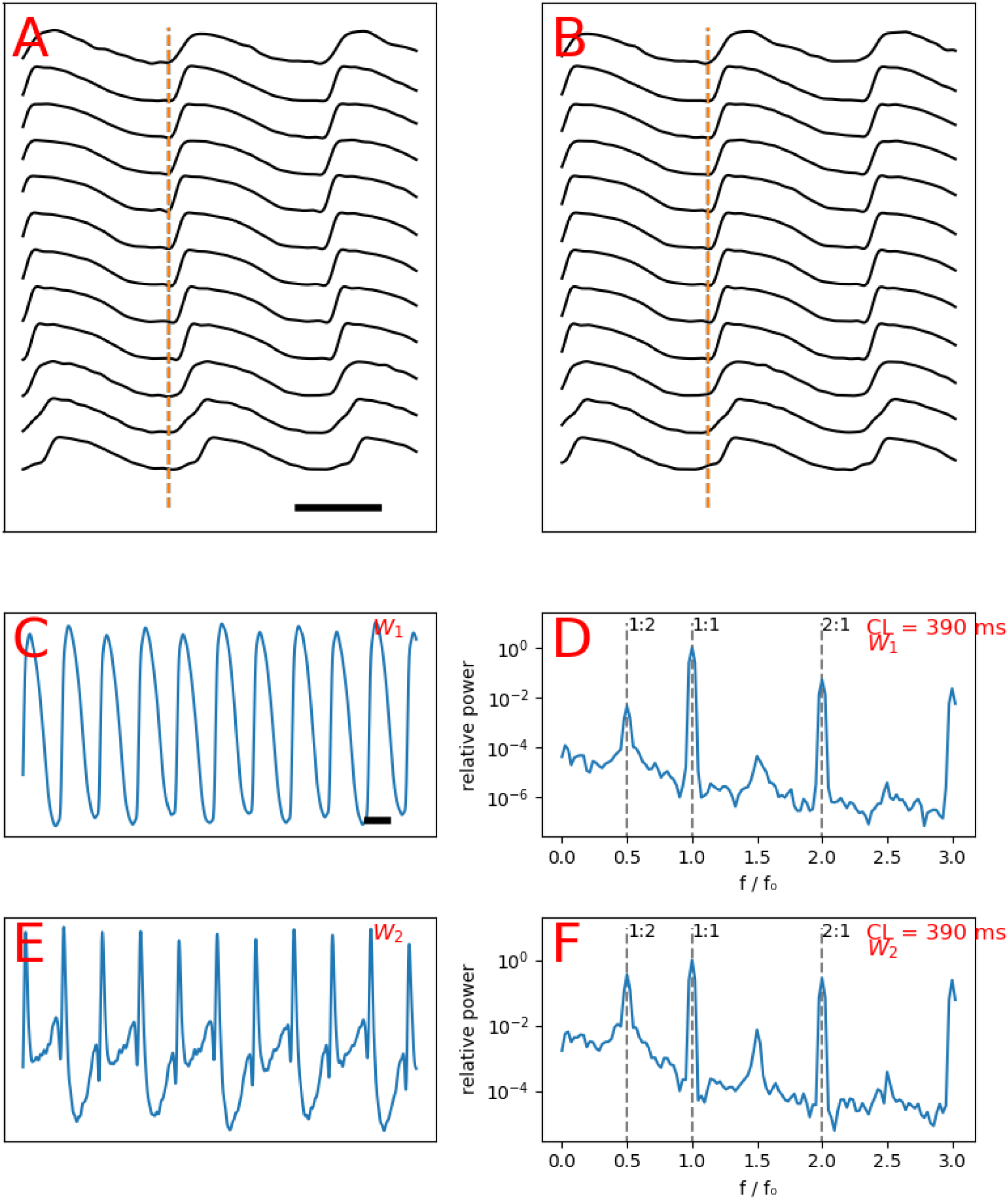
The schematics of global analysis. Spatiotemporally processed signals recorded from multiple points on a line, showing staggered action potentials upstrokes consistent with wavefront propagation (**A**). Same signals as A shifted to align the upstrokes (**B**). The first principle component (*W*_1_) displays alternans (**C**). Spectrogram of C, showing a 1:2 peak of alternans (**D**). The second principle component (*W*_2_) displays more pronounced alternans larger than C (**E**). Spectrogram of E, shows a prominent 1:2 peak of alternans (**F**). The bar in A depicts 200 ms.

In addition to global analysis, we apply local analysis to find the dominant periodicity (i.e., the most common period) of each pixel expressed as an integer in the range 1 to 8. The local analysis uses a combinatorial algorithm, which is described in Supplement C.^22^

### Restitution Curve

According to the restitution hypothesis, the APD is a function of the preceding diastolic interval (DI). The local electrophysiological characteristics of each pixel are evaluated by calculating the local restitution curve by fitting an exponential curve to the (DI, APD) data points obtained from pacing the heart at different cycle lengths (Figure 2A). The exponential curve is parametrized as,

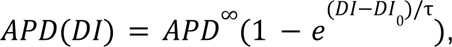

**Figure 2.**
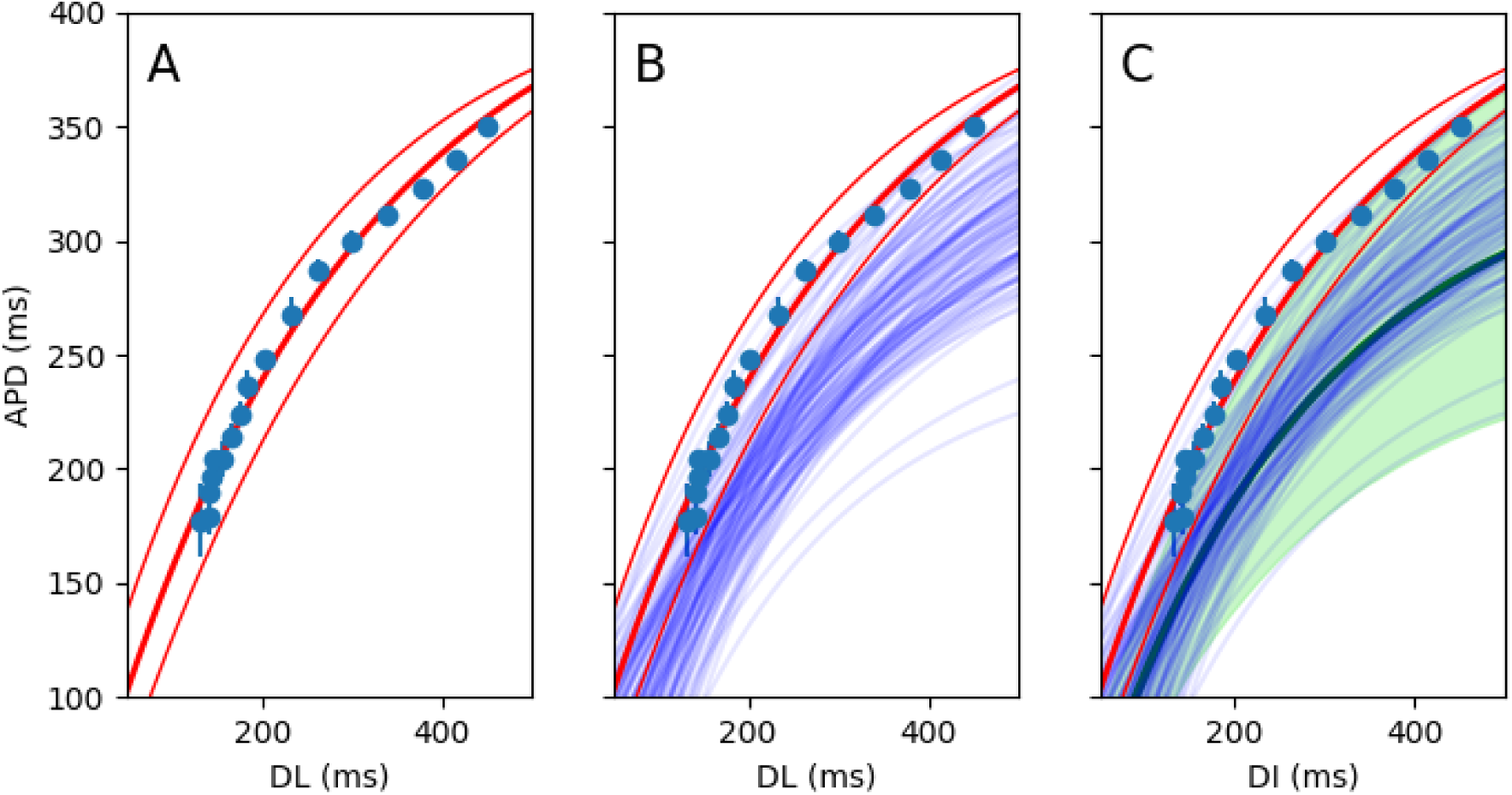
The schematic of composite restitution curve generation. An exponential curve fits a set of (DI, APD) data points for one pixel. The 95% confidence-interval curves are also shown (**A**). The restitution curves for all the pixels in a region of interest (say, with period-2 on the local analysis) are calculated (only 100 curves are shown here)(**B**). Monte Carlo sampling is performed to generate a composite restitution curve (the thick green line) and the confidence interval (shown in a green shade)(**C**).

**Figure 3.**
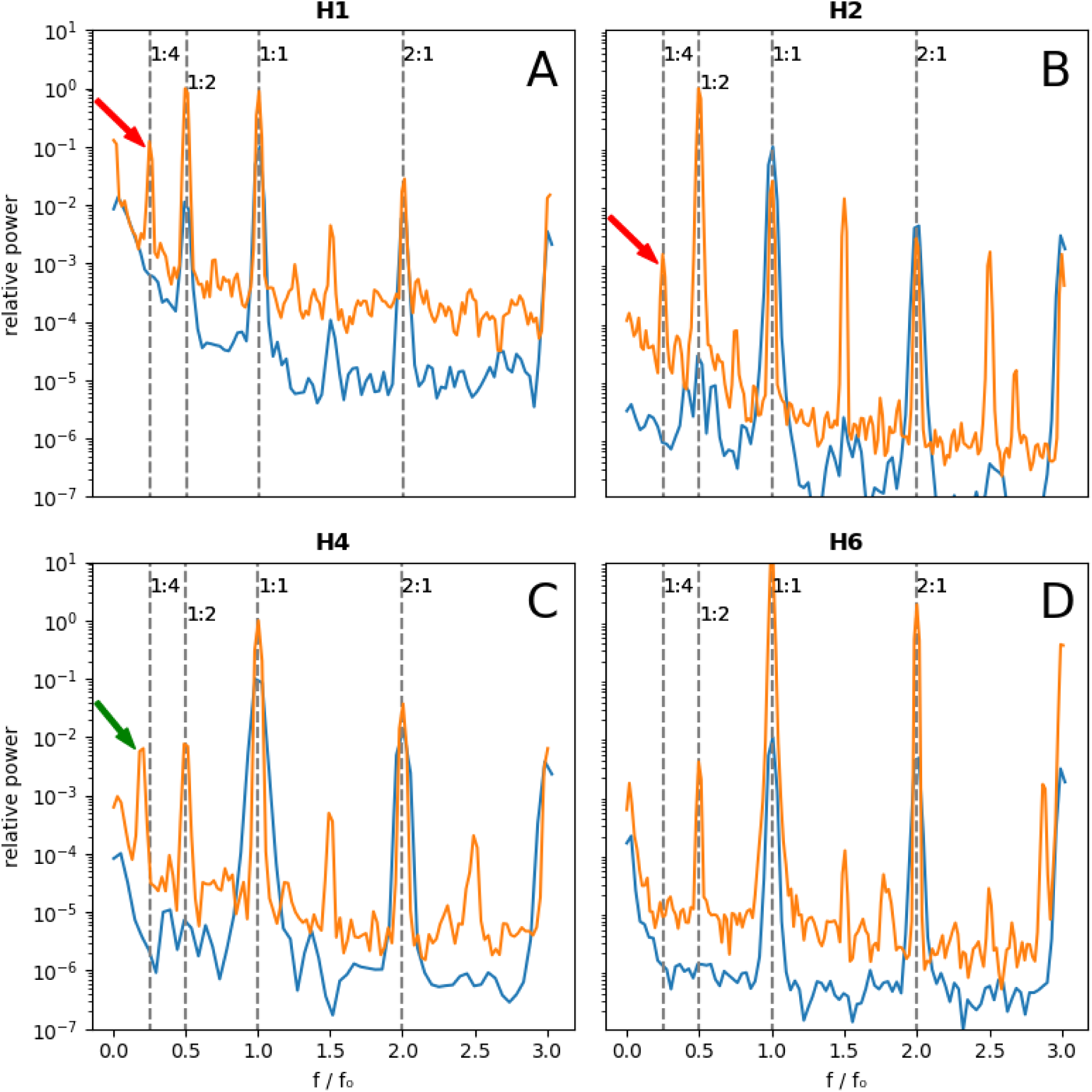
Comparison of baseline and pre-VF spectrograms using global analysis. The blue spectrograms are the baseline (stimulation cycle length of 500 ms except for **H4** at 800 ms) and the orange spectrograms are obtained just before VF induction. **H1**, **H2** (**A** and **B**) exhibit prominent 1:4 peaks (the red arrows), while no discernable 1:4 peak is seen for H6 (**D**). **H4** has a ∼0.18 peak (the green arrow), corresponding to mainly period-6 activity **(C)**. The baseline signals are multiplied by 0.1 to offset the signals for better visualization.

where *APC*^∞^ is the APD steady-state achieved at long cycle lengths, *DI*_0_ is the interception of the exponential curve and the x-axis, and τ is the time-constant measuring the steepness of the restitution curve. Figure 2 (panels B and C) shows how to combine multiple restitution curves to derive a composite curve with confidence intervals.

### Conduction Velocity

Pixel-wise conduction velocity is calculated from the smoothed activation map, i.e., the map showing the time of the upstroke of action potentials for each pixel. Specifically, the conduction velocity is inversely proportional to the magnitude of the gradient of the activation time.

## Results

### The General Characteristics of the Hearts

We report on six explanted human hearts (designated **H1** to **H6**) removed during heart transplantation surgery. Demographics and clinical history of the hearts are presented in Table 1. The aggregate electrophysiological properties of each heart, as described by the restitution curve parameters, are presented in Table 2.

**Table 1:**
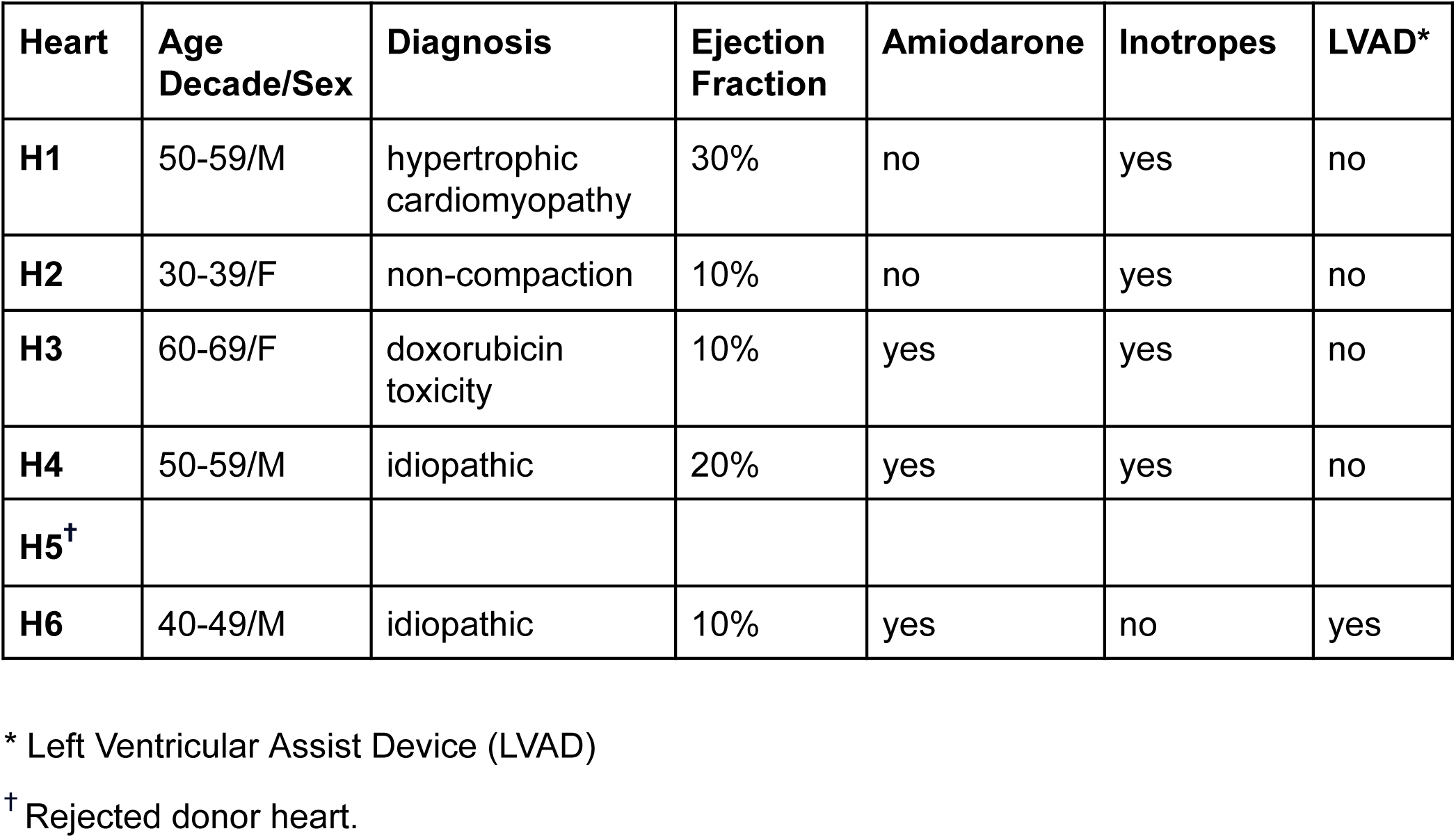
the baseline characteristics of the hearts.

**Table 2:** the electrophysiological characteristics (restitution-curve parameters) of the hearts.

| Heart | $APD^{\infty}$ | $DI_0$ | $\tau$ |
| --- | --- | --- | --- |
| H1 | $383 \pm 43$ | $-18 \pm 52$ | $302 \pm 79$ |
| H2 | $300 \pm 26$ | $-84 \pm 42$ | $318 \pm 65$ |
| H3 | $289 \pm 30$ | $111 \pm 59$ | $69 \pm 47$ |
| H4 | $351 \pm 29$ | $-72 \pm 40$ | $421 \pm 68$ |
| H5 | $261 \pm 71$ | $124 \pm 90$ | $120 \pm 100$ |
| H6 | $419 \pm 24$ | $-51 \pm 17$ | $374 \pm 44$ |

### Global Analysis Detects 1:4 Peaks

Three hearts (**H1**, **H2**, and **H3**) showed a prominent and statistically significant (> 3σ) 1:4 peak. A borderline peak of uncertain significance was seen in another one (**H4**). No 1:4 peak was present in two hearts (**H5** and **H6**).

Figure 2 (panel A) depicts the global spectrogram of **H1**. The baseline spectrogram at 500 ms shows the expected 1:1 peak (the primary activation) and a small 1:2 peak, signifying repolarization alternans. As the heart was stimulated faster at a cycle length of 310 ms, the 1:2 peak became larger, and a new 1:4 peak emerged. This means that a bifurcation occurred somewhere between 500 ms and 310 ms, and the global dynamics had period-4 periodicity. Pacing this heart faster at 300 ms resulted in VF.

**H2** follows a similar pattern (panel B). Again, we observed barely discernible alternans at 500 ms with a strong 1:2 peak and a clear 1:4 peak at 270 ms. Similarly, VF was induced while pacing at 260 ms.

The peak of interest in **H5** (panel C) is offset from the 1:4 location and is around ∼0.18. This peak is further discussed below.

**H6** is a negative example with no 1:4 peak (panel D). There is a prominent 1:2 alternans peak while stimulating at 270 ms, but no significant peak at 1:4. Stimulating this heart faster resulted in a conduction block but no reentrant arrhythmia.

The absence of the 1:4 peak at 500 ms in hearts with a prominent 1:4 peak at short cycle lengths significantly reduces the chance that this peak is a processing artifact and points to its dynamic origin. We can probe the dynamics further by looking at the stimulation frequency dependencies of the 1:2 and 1:4 peaks (Figure 4).

**Figure 4.**
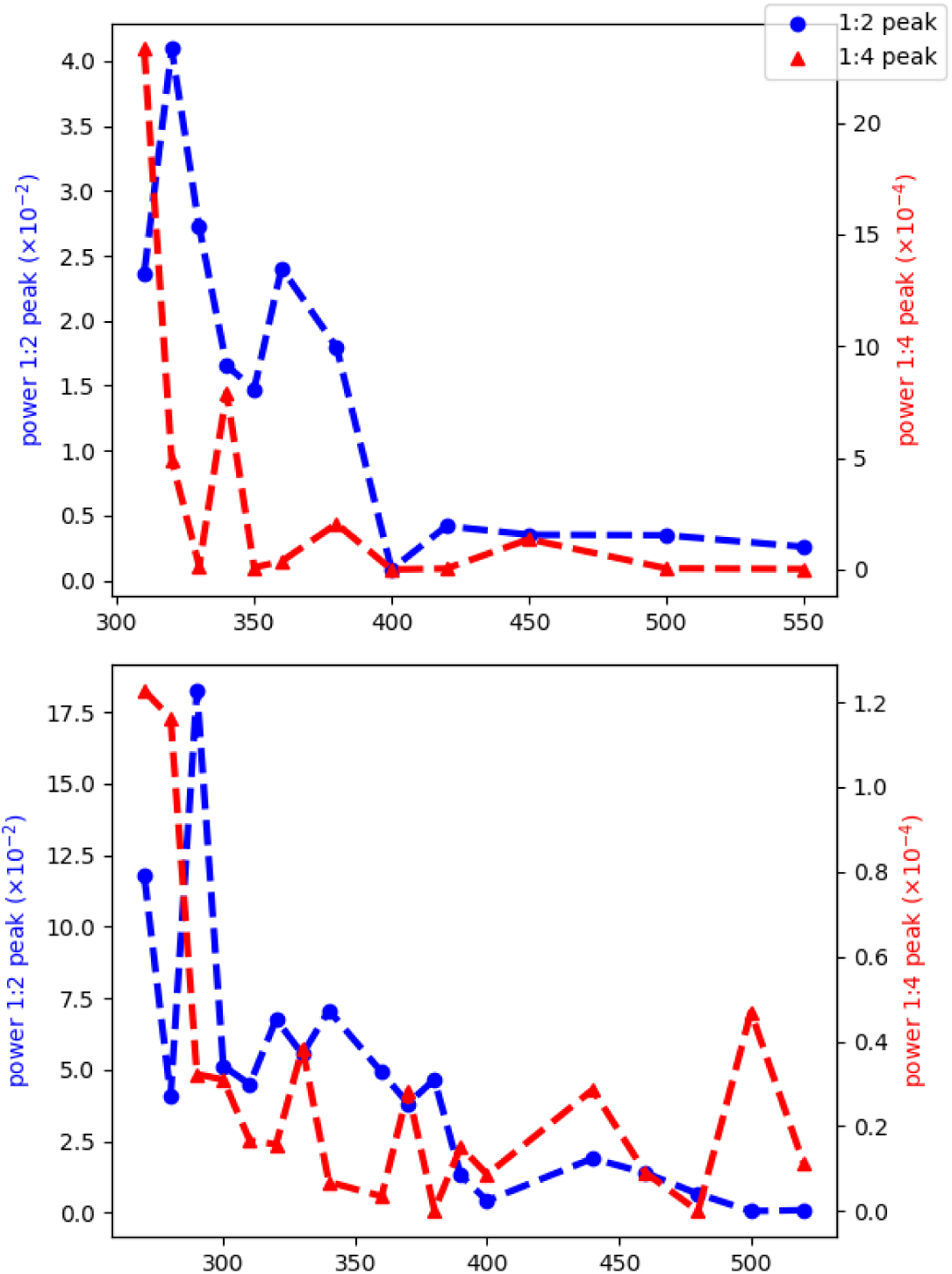
The pacing frequency dependence of 1:2 and 1:4 peaks. In both panels, the 1:2 peak (the classic APD alternans) starts when the cycle length decreases to ∼400 ms. On the other hand, the 1:4 peak only rises above the baseline once the cycle length decreases to 300-350 ms. Also, note the different scaling of the 1:4 peak compared to the 1:2 peak, which is 2-3 orders of magnitude smaller than the 1:2 peak. These results significantly reduce the chance that the observed 1:4 peaks are processing artifacts and point to their dynamical origin.

### Visualizing Higher-Periodicity Signals

Figure 5 lists representative examples of period-2, period-4, period-6, period-8, and higher-order chaotic signals and the corresponding APD trends. Period-4 is stable, but higher-order periods are intermittent. Specifically, the period-8 signal (Figure 5G/H) is irregular but still has sufficient periodicity to be annotated period-8 by the local algorithm.

**Figure 5.**
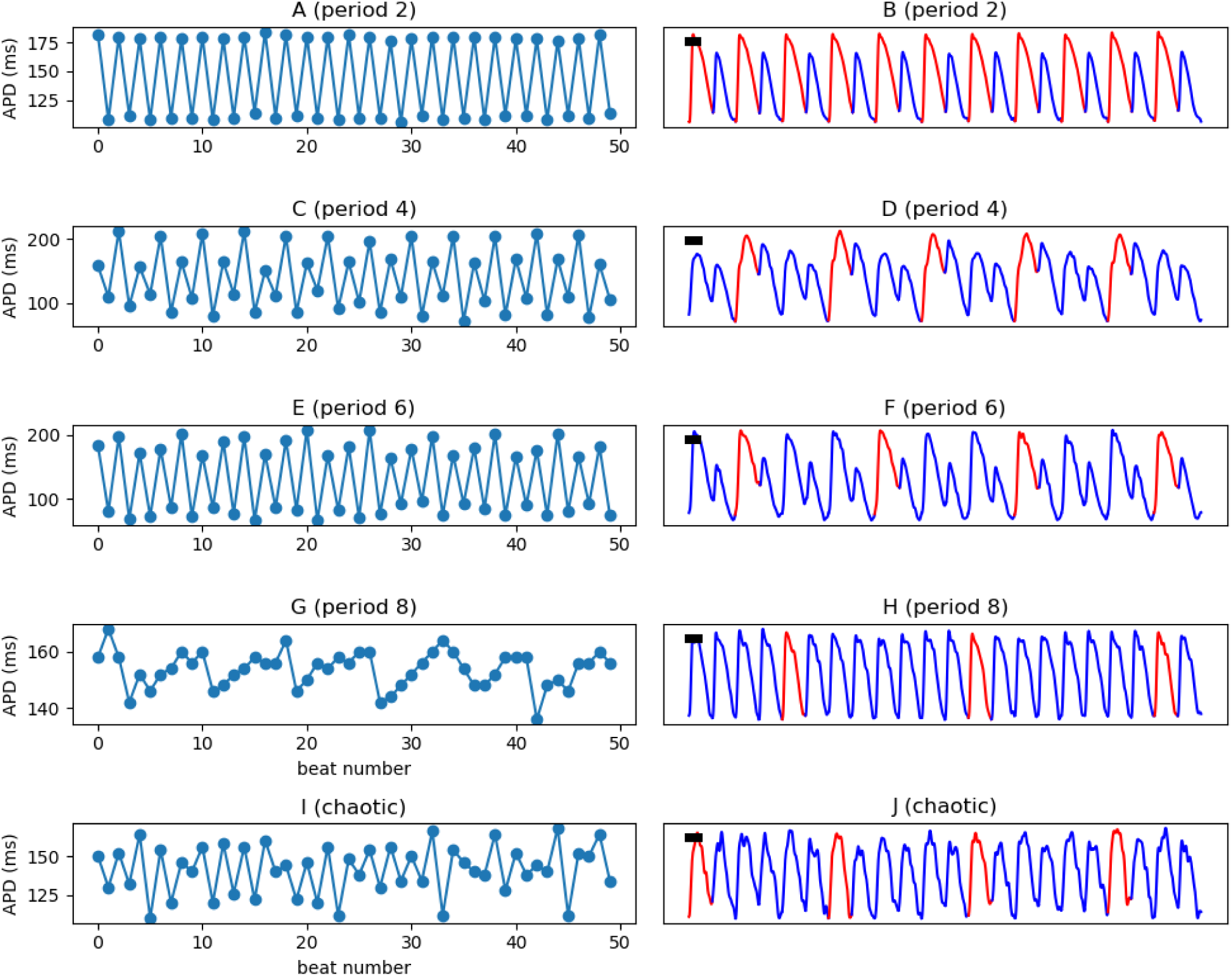
Examples of signals with different periodicities. Representative pixel-level optical-mapping signals with period-2 (**A**), period-4 (**B**), period-6 (**C**), period-8 (**D**), and higher-order/chaotic (**E**) are shown. Note the intermittency of period-8.

### Local Analysis Detects both Period-4 and Higher

The distribution of areas with higher-order periodicity is heterogeneous in both space and time and is variable in different hearts. The spatial distribution of the dominant periodicity in four hearts is depicted in Figure 6.

**Figure 6.**
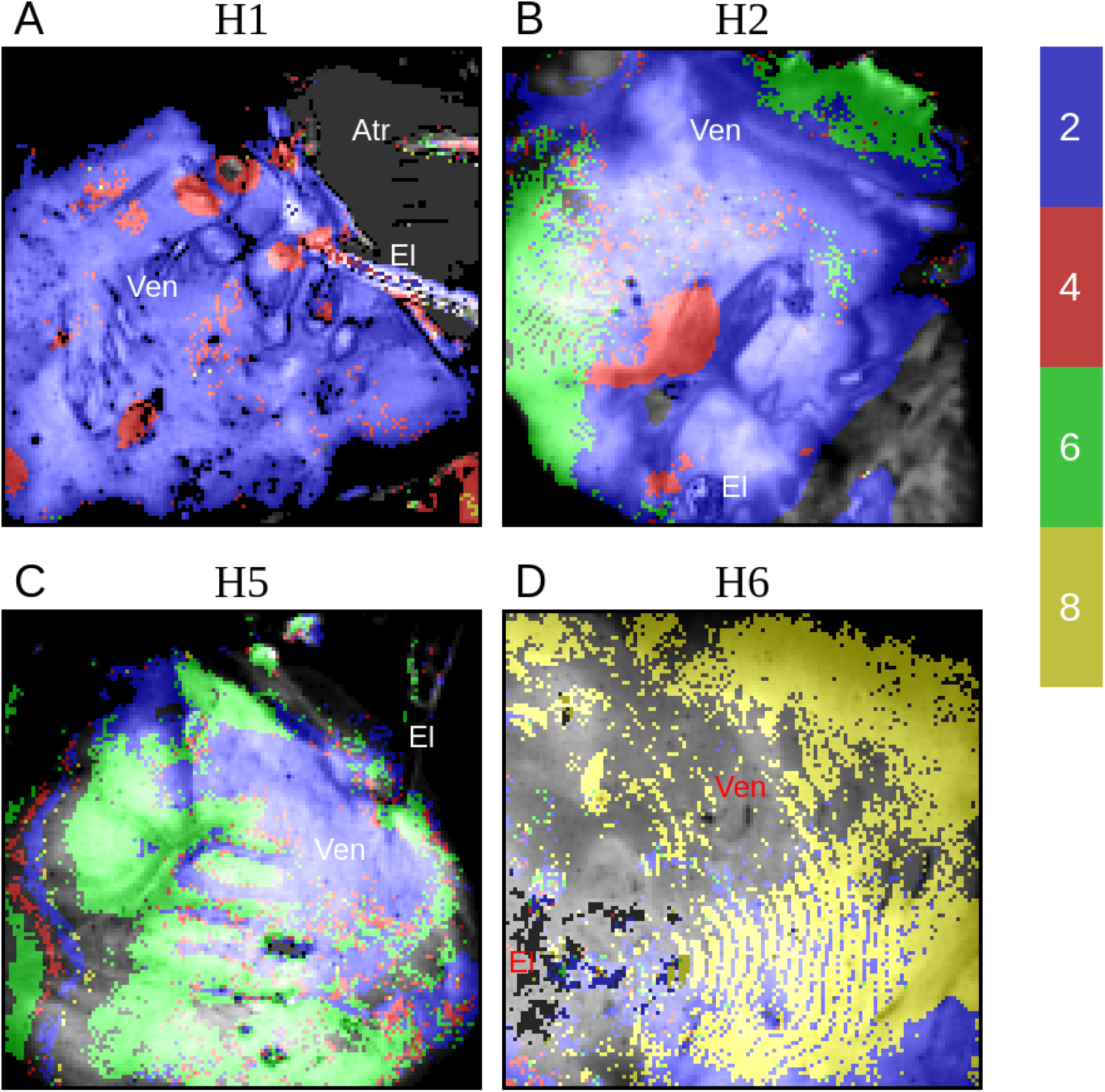
The distribution of higher-order areas. In **H1**, there are multiple period-4 areas (red) in a sea of classic alternans (blue)(**A**). **H2** shows both 1:4 and 1:6 (green) areas (**B**). **H5** has large areas of period-6 without significant period-4 (**C**). The main feature of **H6** is a large area of period-8 without significant 1:2 or 1:4 regions (**D**). The background images (gray-colored) represent anatomy. **Atr**, the atrial; **Ven**, the ventricles; **El**, the pacing electrode.

**H1**, which shows a dominant 1:4 peak in global analysis, has areas of stable period-4 periodicity localized to a few discrete islands with roughly circular borders (Figure 6A). The rest of the ventricle exhibits period-2 with no significant period-6 or higher. On the other hand, **H5**, which does not have an obvious 1:4 peak, has large areas of 1:6 periodicity with no significant amount of 1:2 periodicity (Figure 6C). **H2** is a mixed case with both islands of 1:2 and large areas of 1:6 periodicity (Figure 6B). **H6** is unusual in having large regions of period-8 (Figure 5D); however, it should be noted that **H6** is the only heart in our study that was on a left ventricular assist device (LVAD) before transplantation (Table 1).

Figure 7 displays the relative proportion of pixels of different periodicities in the six study hearts. Again, we noted the significant variability among the hearts. The results are consistent with the global analysis. For example, **H1**, **H2**, and **H3** have clear 1:4 peaks in global analysis and a significant period-4 peak in the histograms, whereas **H6** has no 1:4 peak in global analysis and a barely discernible period-4 peak in the histogram.

**Figure 7.**
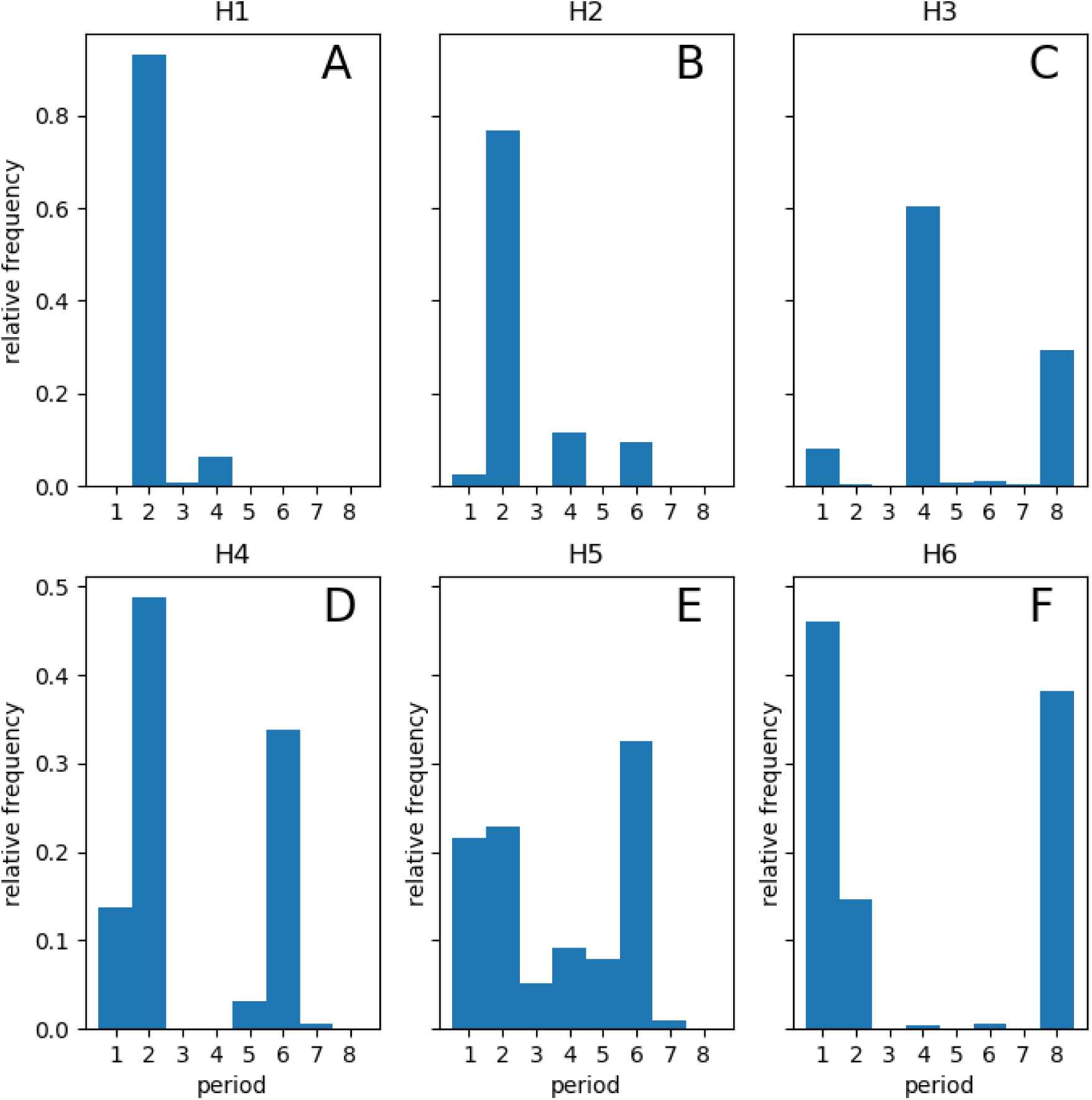
The relative frequency of different periodicities. Each histogram shows the relative proportions of pixels with a given period (in the range 1 to 8) for each of the six hearts.

### Effects of Amiodarone

Three hearts (**H3**, **H4**, and **H6**) were on amiodarone, a multi-channel membrane active antiarrhythmic medication (Table 1). According to the histograms in Figure 7, the main effect of amiodarone is to promote the 1:1 peak, i.e., to increase the size of the areas without alternans.

### Electrophysiological Characteristics of Higher-Order Areas

Figure 8 compares the electrophysiological properties between areas of different periodicity in the same heart. For each heart, the aggregate restitution curve and conduction velocity of the period-2 regions is used as control and is compared to the aggregate properties of the regions with period-4, -6, or -8, as long as the area is large enough to show an obvious peak in the histograms.

**Figure 8.**
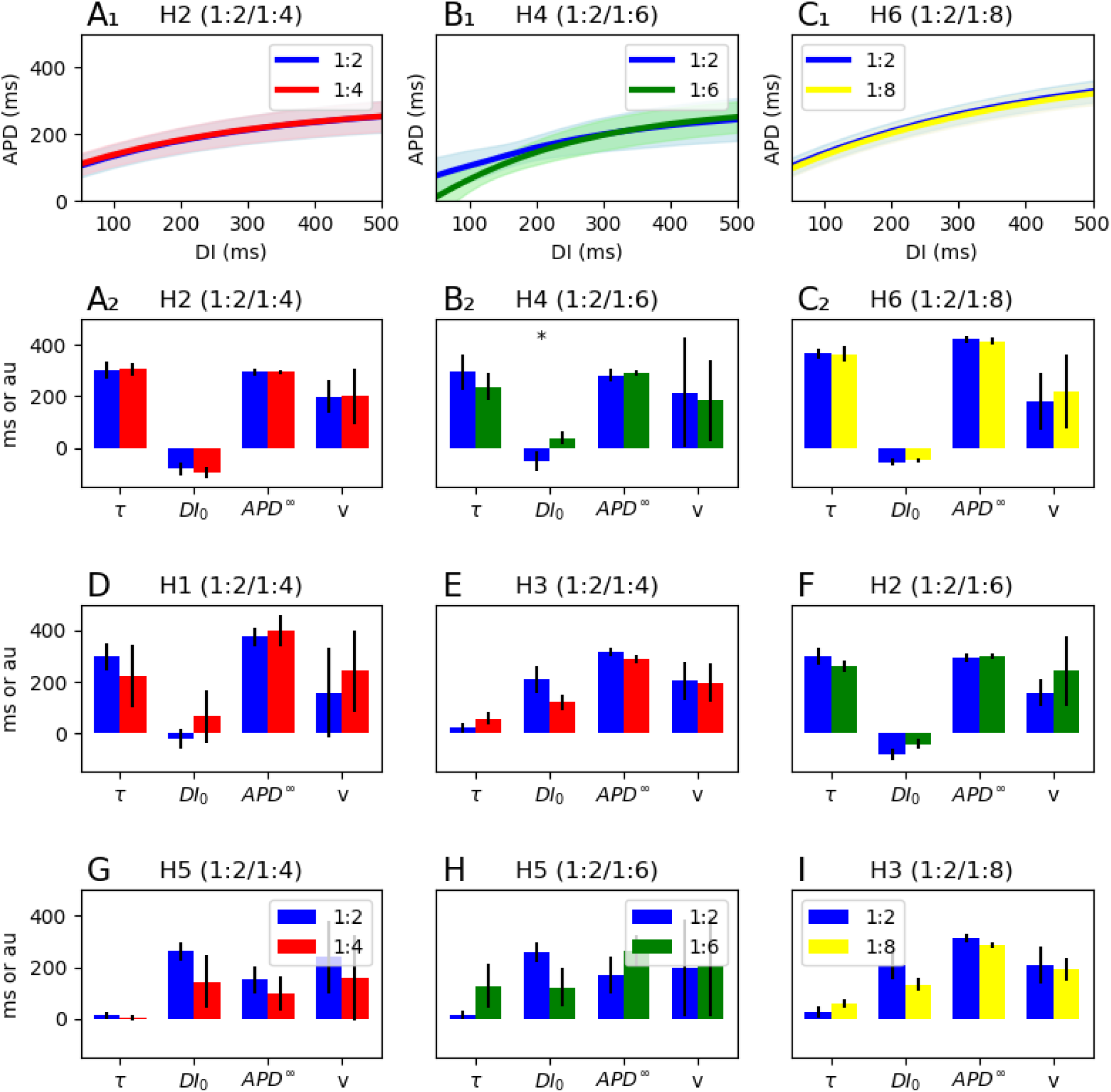
The electrophysiological characteristics of areas with different periodicities. The top row (**A1**, **B1**, **C1**) compares restitution curves, whereas the other panels show the electrophysiological properties (restitution curve parameters and conduction velocity) between different periodicities. Each panel applies to one heart. Only one comparison (B2) shows any significant difference in the baseline electrophysiological properties, with a marginal separation in the corresponding restitution curves (B1).

There is no significant difference in the electrophysiological characteristics of the 1:2, 1:4, and 1:8 areas, which are essentially indistinguishable. Only one of the three 1:2 vs. 1:6 comparisons shows a marginal difference (Figure 8B1/B2). In other words, *the underlying electrophysiological properties, as measured by the restitution curve and local conduction velocity at slower pacing rates, is not a predictor of the dynamics when pacing at faster rates*.

## Discussion

We report what we believe to be the first detection of stable period-4 and intermittent period-6, and period-8 in human cardiac tissue during fast stimulation. There is a large degree of heterogeneity in the distribution of higher-order periodic areas among the hearts and in different regions of the same heart. However, periodicities larger than 2 (i.e., beyond classic alternans) were detected in all six hearts, and they are likely a common occurrence in severely diseased human hearts at sufficiently fast pacing rates. Moreover, we observed the coexistence of higher-period regions with lower-period regions (period-1 and period-2); which suggests there is a period of time when portions of the ventricles are already electrically unstable, but the entire heart is not in VT or VF yet.

We observed no significant baseline electrophysiological differences between areas with 1:2 periodicity and areas with period-4, -6, and -8. Specifically, restitution curves remain the same. Therefore, the baseline restitution properties do not predict the ultimate pre-VF dynamics.

These results have mechanistic implications. There are two known mechanisms for generating alternans and higher-order periodicities: voltage-driven (dependent on the restitution properties) and calcium-driven.^14,23^ Based on the observed independence of the local dominant period from the restitution properties, we infer that the oscillation of the calcium machinery and excitation-contraction coupling is the primary driver of higher-order dynamics in human hearts. This result is consistent with theoretical works that showed monotonically increasing restitution curves, as is the case for human hearts, cannot generate periods greater than 2.^24^

According to the concordant to discordant alternans pathway, the heart is in a meta-stable but not chaotic condition just before VF initiation, waiting for a trigger. The trigger is usually conduction block or a premature ventricular contraction caused by early after depolarization (EAD) or phase-2 reentry.^25,26^ Higher-order and chaotic regions may spatially grow to act as niduses of instability that then degenerate regular rhythms into chaotic fibrillation.

As mentioned prior, one of the main motivations behind this study was to develop a theoretical framework for possible substrate-directed ablation of VF. Our results, as relevant to VF ablation, are mixed. On the one hand, detecting higher-order periodicities in human hearts, the spatial heterogeneity of these regions, and the co-existence of chaotic pockets and normal rhythm, are promising evidence that target regions exist. On the other hand, we noted that the restitution properties and conduction velocity are not predictors of these areas; therefore, it is unclear how we can locate these targets during ablation. Our results do not preclude the possibility that local differences in calcium cycling properties can predict the site of higher-order dynamics; however, no clinically feasible technique exists to measure these characteristics.

We did observe larger non-alternating areas (period-1) in amiodarone-treated hearts relative to higher-order periodicities, which points to the membrane-stabilizing effects of amiodarone. Nevertheless, this effect is partial, and the amiodarone-treated hearts still exhibit higher-order periodicities.

Detecting higher-order periodicities in human hearts may have practical implications beyond its relevance to ablation. If a practical method to detect period-4 from clinical recordings (e.g., surface ECG) is developed, it might fix the main shortcoming of T wave/APD alternans in the form of low positive predictive value for malignant ventricular arrhythmias.^27^

### Limitations

Human recipient hearts used in this study are very heterogeneous with variable age, sex, underlying disease, pre-transplant ejection fraction, exposure to different antiarrhythmic medications, and the use of mechanical circulatory support before transplantation. In addition, the logistical problems in performing the experiments prevent using a large sample size. The combination of the small sample size and heterogeneous hearts makes it difficult to perform accurate statistical analysis. While the primary finding of the study, i.e., the detection of higher-order periodicities in human hearts, is robust, the secondary findings, primarily as related to the mechanistic differences, are less confident and should be considered mainly as hypothesis generation rather than confirmatory. In addition, we cannot prospectively control for confounding factors, like exposure to amiodarone and inotropes.

Transplant recipient hearts used in this study are very diseased and electrophysiologically different from normal human hearts. Therefore, the results obtained here may not apply to normal hearts. Nevertheless, these are precisely the hearts prone to malignant arrhythmias that would benefit from ablation. The results obtained from recipients’ hearts are likely more clinically relevant than studies done on normal hearts.

## Conclusion

Focal areas of higher-order periods can occur in diseased human hearts under fast pacing. The underlying electrophysiological characteristics, as measured by the restitution properties and conduction velocity, are not predictive of the localization of these areas.

## Data Availability

The de-identified processed data products, analysis results, and software code are available upon request.

## Acknowledgments

The study protocol was approved by the Emory University and Georgia Institute of Technology Institutional Review Boards (IRB) under Protocol Number H22204.

## References

1. Weiss JN, Qu Z, Chen P-S, Lin S-F, Karagueuzian HS, Hayashi H, Garfinkel A, Karma A. The dynamics of cardiac fibrillation. Circulation. 2005;112:1232–1240.

2. Fenton FH, Cherry EM, Hastings HM, Evans SJ. Multiple mechanisms of spiral wave breakup in a model of cardiac electrical activity. Chaos Woodbury N. 2002;12:852–892.

3. Jalife J. Ventricular Fibrillation: Mechanisms of Initiation and Maintenance. Annu. Rev. Physiol. 2000;62:25–50.

4. Al-Khatib SM, Stevenson WG, Ackerman MJ, Bryant WJ, Callans DJ, Curtis AB, Deal BJ, Dickfeld T, Field ME, Fonarow GC, et al. 2017 AHA/ACC/HRS Guideline for Management of Patients With Ventricular Arrhythmias and the Prevention of Sudden Cardiac Death. Circulation. 2018;138:e272–e391.

5. Di BL, Burkhardt JD, Lakkireddy D, Carbucicchio C, Mohanty S, Mohanty P, Trivedi C, Santangeli P, Bai R, Forleo G, et al. Ablation of Stable VTs Versus Substrate Ablation in Ischemic Cardiomyopathy. J. Am. Coll. Cardiol. 2015;66:2872–2882.

6. Tung R, Vaseghi M, Frankel DS, Vergara P, Di Biase L, Nagashima K, Yu R, Vangala S, Tseng C-H, Choi E-K, et al. Freedom from recurrent ventricular tachycardia after catheter ablation is associated with improved survival in patients with structural heart disease: An International VT Ablation Center Collaborative Group study. Heart Rhythm. 2015;12:1997–2007.

7. Aziz Z, Shatz D, Raiman M, Upadhyay GA, Beaser AD, Besser SA, Shatz NA, Fu Z, Jiang R, Nishimura T, et al. Targeted Ablation of Ventricular Tachycardia Guided by Wavefront Discontinuities during Sinus Rhythm: A New Functional Substrate Mapping Strategy. Circulation. 2019;140:1383–1397.

8. Kakihara J, Takagi M, Hayashi Y, Tatsumi H, Doi A, Yoshiyama M. Radiofrequency catheter ablation for treatment of premature ventricular contractions triggering ventricular fibrillation from the right ventricular outflow tract in a patient with early repolarization syndrome. Hear. Case Rep. 2016;2:342–346.

9. Prevention of Ventricular Fibrillation Episodes in Brugada Syndrome by Catheter Ablation Over the Anterior Right Ventricular Outflow Tract Epicardium | Circulation.

10. Haïssaguerre M, Shoda M, Jaïs P, Nogami A, Shah DC, Kautzner J, Arentz T, Kalushe D, Lamaison D, Griffith M, et al. Mapping and Ablation of Idiopathic Ventricular Fibrillation. Circulation. 2002;106:962–967.

11. Pastore JM, Girouard SD, Laurita KR, Akar FG, Rosenbaum DS. Mechanism linking T-wave alternans to the genesis of cardiac fibrillation. Circulation. 1999;99:1385–1394.

12. Pastore JM, Rosenbaum DS. Role of structural barriers in the mechanism of alternans-induced reentry. Circ. Res. 2000;87:1157–1163.

13. Narayan SM. T-Wave Alternans and the Susceptibility to Ventricular Arrhythmias. J. Am. Coll. Cardiol. 2006;47:269–281.

14. Kulkarni K, Merchant FM, Kassab MB, Sana F, Moazzami K, Sayadi O, Singh JP, Heist EK, Armoundas AA. Cardiac Alternans: Mechanisms and Clinical Utility in Arrhythmia Prevention. J. Am. Heart Assoc. 2019;8:e013750.

15. Gizzi A, Cherry EM, Gilmour RF, Luther S, Filippi S, Fenton FH. Effects of pacing site and stimulation history on alternans dynamics and the development of complex spatiotemporal patterns in cardiac tissue. Front. Physiol. 2013;4.

16. Savino GV, Romanelli L, González DL, Piro O, Valentinuzzi ME. Evidence for chaotic behavior in driven ventricles. Biophys. J. 1989;56:273–280.

17. Gilmour RF, Otani NF, Watanabe MA. Memory and complex dynamics in cardiac Purkinje fibers. Am. J. Physiol. [Internet]. 1997 [cited 2023 Jan 11];272. Available from: https://pubmed.ncbi.nlm.nih.gov/9139969/

18. Fedorov VV, Lozinsky IT, Sosunov EA, Anyukhovsky EP, Rosen MR, Balke CW, Efimov IR. Application of blebbistatin as an excitation–contraction uncoupler for electrophysiologic study of rat and rabbit hearts. Heart Rhythm. 2007;4:619–626.

19. Efimov IR, Nikolski VP, Salama G. Optical imaging of the heart. Circ. Res. 2004;95:21–33.

20. Matiukas A, Mitrea BG, Qin M, Pertsov AM, Shvedko AG, Warren MD, Zaitsev AV, Wuskell JP, Wei M, Watras J, et al. Near-infrared voltage-sensitive fluorescent dyes optimized for optical mapping in blood-perfused myocardium. Heart Rhythm. 2007;4:1441–1451.

21. Uzelac I, Iravanian S, Bhatia NK, Fenton FH. Direct observation of a stable spiral wave reentry in ventricles of a whole human heart using optical mapping for voltage and calcium. Heart Rhythm. 2022;19:1912–1913.

22. Iravanian S. Weakly-Coupled Oscillators with Long-Distance Correlation as a Model of Human Atrial Fibrillation. bioRxiv. 2021;

23. Qu Z, Weiss JN. Cardiac Alternans: From Bedside to Bench and Back. Circ. Res.2023;132:127–149.

24. Qu Z, Shiferaw Y, Weiss JN. Nonlinear dynamics of cardiac excitation-contraction coupling: an iterated map study. Phys. Rev. E Stat. Nonlin. Soft Matter Phys. 2007;75.

25. Yan G-X, Joshi A, Guo D, Hlaing T, Martin J, Xu X, Kowey PR. Phase 2 Reentry as a Trigger to Initiate Ventricular Fibrillation During Early Acute Myocardial Ischemia. Circulation. 2004;110:1036–1041.

26. Fenton FH, Kim TY, Cherry EM, Uzelac I, Iravanian S, Cho HC, Chionuma H, Shah AD, Burke M, Merchant FM, et al. PO-691-05 First Experimental Observation Of Alternans-Induced Phase-2 Reentry In Brugada Syndrome By Optical Mapping On An Explanted Human Heart, With Numerical Simulation Validation. Heart Rhythm. 2022;19:S400.

27. Gehi AK, Stein RH, Metz LD, Gomes JA. Microvolt T-Wave Alternans for the Risk Stratification of Ventricular Tachyarrhythmic Events: A Meta-Analysis. J. Am. Coll. Cardiol. 2005;46:75–82.

